# Mapping county-level tick-borne disease risk in Illinois by integrating vector abundance, pathogen prevalence, and phenology

**DOI:** 10.64898/2026.10.14.26365569

**Authors:** Abrar Hussain, Suman Bhowmick, Nohra Mateus-Pinilla, Rebecca L. Smith

**Affiliations:** Department of Pathobiology, College of Veterinary Medicine, University of Illinois, Urbana-Champaign, 61802, IL, USA; Illinois Natural History Survey - Prairie Research Institute, University of Illinois Urbana-Champaign, 61802, IL, USA; Department of Biomedical and Translational Sciences, Carle-Illinois College of Medicine, University of Illinois Urbana-Champaign, 61802, IL, USA

**Keywords:** Ticks, Tick-borne disease, *Amblyomma americanum*, *Dermacentor variabilis*, *Ixodes scapularis*, spatial epidemiology, seasonal risk, Illinois, R Shiny

## Abstract

Tick-borne disease risk reflects the combined influence of vector abundance, pathogen prevalence, human exposure, and seasonal activity, yet these components are rarely integrated at the county level. We developed species-specific ecological risk scores for three medically important tick species (Acari: Ixodidae): *Amblyomma americanum* (L.), *Dermacentor variabilis* (Say), and *Ixodes scapularis* Say, across all 102 Illinois counties using data from 2019 to 2022. County-level predicted tick abundance and pathogen prevalence were scaled and combined using equal weights to generate primary vector-level risk scores. Robustness to the equal-weight assumption was evaluated by systematically varying the relative weights assigned to abundance and pathogen prevalence, while a separate sensitivity analysis incorporated reported tick-bite incidence where available. Associations with human disease were evaluated using negative binomial regression of tick-borne disease case counts on the corresponding risk scores, with county population included as an offset. Monthly phenology indicators were constructed by integrating human disease cases, reported tick bites, and sampled tick abundance and were evaluated in relation to temperature and precipitation. Seasonal risk was estimated by combining county-level vector risk with species-specific monthly phenology, generating monthly risk surfaces that were deployed through an interactive R Shiny application. Higher risk scores were significantly associated with greater disease case rates for each tick species. County rankings were highly stable under moderate changes in abundance and prevalence weighting. Seasonal activity differed among species, with *A. americanum* and *D. variabilis* showing concentrated warm-season risk and *I. scapularis* exhibiting a broader seasonal profile. These county-level seasonal risk maps identify where and when relative tick-borne disease risk is elevated in Illinois, supporting targeted prevention, surveillance, and clinical awareness.

## Introduction

Tick-borne diseases are an increasing public health concern in the United States, with reported vector-borne disease cases rising substantially over recent decades (Rosenberg et al., 2019). This increase has coincided with the emergence of additional tick-borne pathogens (R. J. Eisen et al., 2017) and the geographic expansion of several medically important tick species (Sonenshine, 2018). *Ixodes scapularis* Say (Acari: Ixodidae) has expanded across large portions of the northern and central United States (R. J. Eisen et al., 2016), while *Amblyomma americanum* (L.) has extended beyond its historically recognized range (Raghavan et al., 2019; Springer et al., 2014). Geographic expansion of these vectors has also contributed to changes in the distribution of human tick-borne diseases, including Lyme disease (Kugeler et al., 2015). Climate and environmental conditions influence tick survival, development, host-seeking activity, and establishment, further affecting where and when exposure may occur (Brownstein et al., 2003; Deshpande et al., 2024). These changes have increased the need for surveillance approaches that move beyond describing tick occurrence and instead integrate multiple components relevant to human disease risk.

Illinois is an important setting for such an approach because three medically important tick species, *A. americanum, Dermacentor variabilis* (Say), and *I. scapularis*, occur across the state but differ in their spatial distributions and environmental associations. Habitat suitability modeling has identified substantial areas of Illinois as suitable for these species and suggested that suitability may shift under changing climatic conditions (Kopsco et al., 2023). Field-based studies have also demonstrated associations between weather and the abundance of medically important ticks in Illinois (Bacon et al., 2022). Statewide surveillance has identified distinct distribution and clustering patterns among these species (Hussain et al., 2025), while our previous Bayesian spatial models generated county-level predicted abundance estimates for *A. americanum, D. variabilis*, and *I. scapularis* across Illinois (Hussain et al., 2026). Human surveillance data provide an additional layer of information, with retrospective analyses showing considerable spatial heterogeneity in associated tick-borne diseases across the state (Chakraborty et al., 2026). These available datasets provide a strong basis for county-level risk assessment, but vector abundance, pathogen surveillance, human exposure, and disease occurrence have generally been evaluated separately.

Existing approaches provide useful foundations for integrating these components. Classical acarological measures have combined vector abundance and pathogen infection to estimate entomological hazard, particularly through measures based on infected *I. scapularis* nymphs (Diuk-Wasser et al., 2012; Mather et al., 1996). Such measures have shown associations with human Lyme disease incidence (Pepin et al., 2012), but most were developed for a single vector-pathogen system. Human exposure is another component of risk that vector surveillance alone does not capture. Integrated ecological and behavioral mapping has therefore been proposed to improve the identification of public health priorities (Bouchard et al., 2018), while community science programs have provided additional information on where and when people encounter ticks (Kache et al., 2023; Nieto et al., 2018). Crowdsourced observations have also been used to characterize tick distribution and seasonality (Cull, 2022). Temporal variation presents an additional challenge because tick activity differs among species and life stages (Burtis et al., 2023; Hamer et al., 2012; Ogden et al., 2018). CDC surveillance guidance similarly emphasizes county-level characterization of tick distribution, pathogen presence and prevalence, host-seeking tick density, and phenology to identify where and when exposure to infected ticks may occur (CDC, 2019, 2020). Although these ecological, exposure, epidemiological, and seasonal data streams are increasingly available, there remains a practical gap in integrating them across multiple tick species within a common spatial and temporal risk framework.

To address this gap, we developed species-specific county-level ecological risk scores for *A. americanum, D. variabilis*, and *I. scapularis* across Illinois by combining predicted tick abundance with tick-borne pathogen prevalence. We evaluated the robustness of the equal-weight formulation by systematically varying the relative contributions of tick abundance and pathogen prevalence and separately evaluated the influence of reported tick-bite exposure on county-level risk rankings. We then assessed the relationship between the primary risk scores and observed human tick-borne disease burden. We also characterized species-specific phenology using sampled tick abundance, reported tick-bite encounters, and human disease cases, and evaluated associations with temperature and precipitation. Finally, county-level ecological risk was combined with monthly phenology to generate seasonal risk estimates, which were incorporated into an interactive R Shiny application. This framework was designed to provide a unified representation of where and when relative tick-borne disease risk is elevated across Illinois while integrating ecological hazard, human exposure, epidemiological outcomes, and seasonality.

## Materials and Methods

### Data curation

County-level data were compiled for all 102 Illinois counties for three medically important tick species: *A. americanum, D. variabilis*, and *I. scapularis*. The datasets included previously estimated county-level tick abundance (Hussain et al., 2026), tick-borne pathogen (TBP) prevalence extracted from the Illinois Department of Public Health (IDPH) (*Illinois Tickborne Disease Dashboard*, n.d.), reported tick-bite encounters from the I-Tick app (*Illinois - The Tick App*, n.d.), human tick-borne disease (TBD) cases from IDPH, and county population data for 2020 (*Illinois - Census Bureau Profile*, n.d.). Tick abundance, tick-borne pathogen prevalence, tick-bite encounters, and Human TBD case data were compiled from cumulative observations from 2019 through 2022 to ensure uniformity. County-level tick distribution was represented by the posterior-mean predicted abundance of all three tick species, obtained from previously developed spatial models (Hussain et al., 2026). Tick-borne pathogen prevalence was calculated separately for *A. americanum, D. variabilis*, and *I. scapularis* in each county by pooling IDPH testing records across 2019-2022 and dividing the cumulative number of pathogen-positive ticks by the cumulative number of ticks tested. Pathogen prevalence was then spatially smoothed to reduce instability associated with limited county-level testing (Supplementary materials). Reported tick-bite encounters were aggregated separately for all three tick species within each county across 2019-2022. The annual average tick-bite incidence per 100,000 population was calculated as the cumulative number of reported encounters for each species. Human TBD cases were grouped according to the principal vector species. Ehrlichiosis cases were assigned to *A. americanum*, spotted fever rickettsioses cases were assigned to *D. variabilis*, and anaplasmosis, babesiosis, and Lyme disease cases were summed and assigned to *I. scapularis*. For each vector-associated disease group, cumulative cases from 2019-2022 were used to calculate the annual average incidence per 100,000 population. County names were standardized across all datasets, and the processed data were merged with the Illinois county boundary shapefile using county as the common geographic identifier. To place variables measured on different scales onto a comparable range, county-level tick abundance, tick-borne pathogen prevalence, and tick-bite incidence were scaled separately for each tick species (Supplementary materials) (Figure 1).

**Figure 1.**
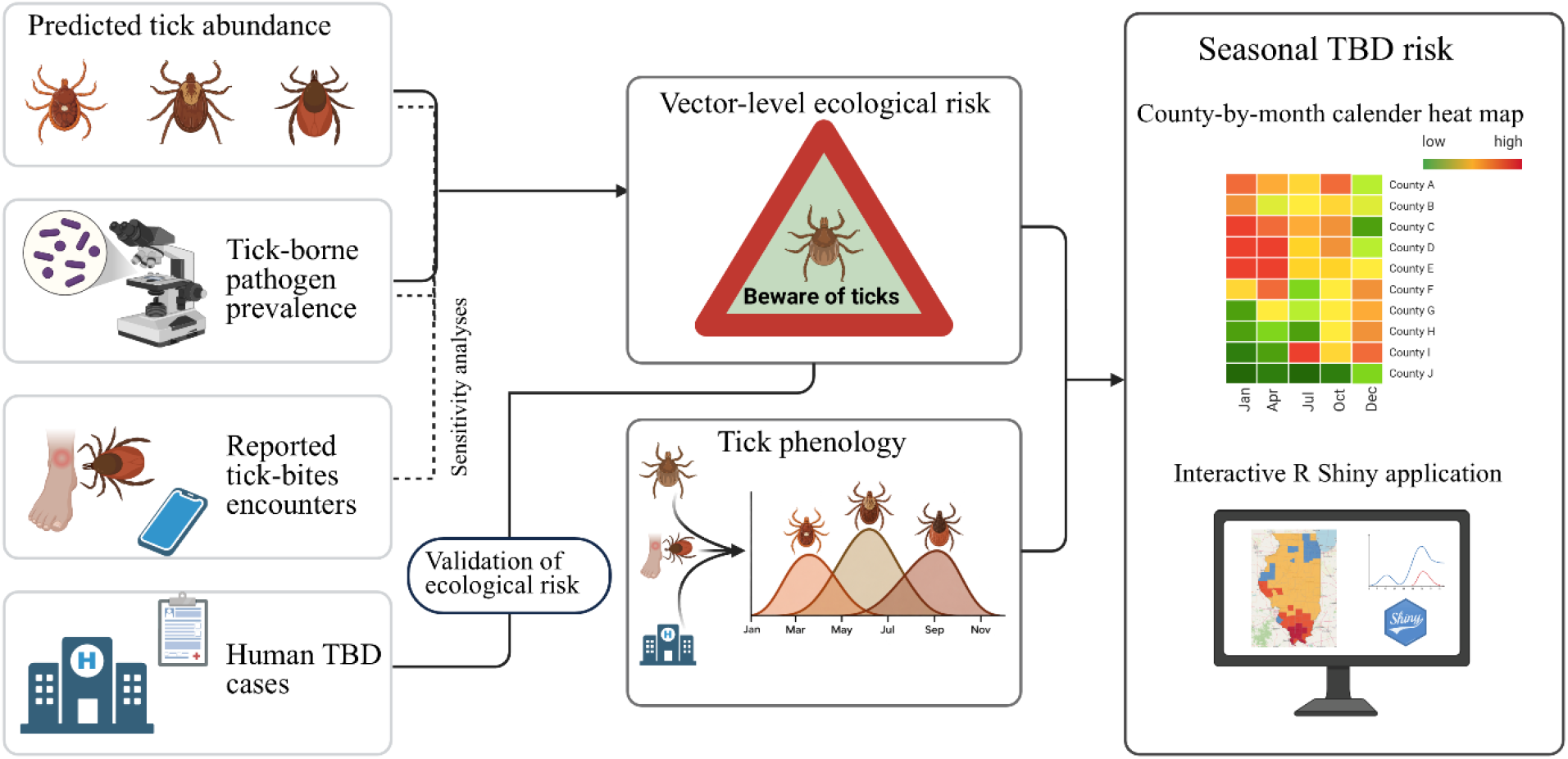
Analytical framework for county-level seasonal tick-borne disease risk in Illinois. Tick abundance and pathogen prevalence were combined into species-specific ecological risk scores, with alternative weighting and tick-bite exposure evaluated through sensitivity analyses. Human disease data were used for validation, and monthly phenology was integrated with ecological risk to generate seasonal risk estimates and an interactive R Shiny application.

### Vector-level risk score construction and sensitivity analyses

A vector-level ecological risk score was constructed for each of the three tick species by combining tick abundance with TBP prevalence. The vector-level risk score was calculated as an equal-weight additive combination:

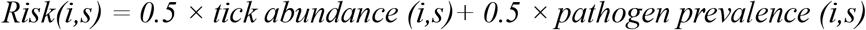

where i denotes the county and s denotes the tick species. Equal weighting was used so that tick abundance and pathogen prevalence contributed equally to the vector-level ecological risk score.

To evaluate the robustness of county-level rankings to the equal-weight assumption, a deterministic weight sensitivity analysis was conducted. The abundance weight varied from 0.00 to 1.00 in increments of 0.05, with the pathogen-prevalence weight constrained to one minus the abundance weight. At each weight combination, species-specific risk scores were recalculated, and county rankings were compared with the primary 50:50 formulation using Spearman’s rank correlation. The highest-scoring 20% of counties, corresponding to 21 of 102 counties, were used as a fixed high-priority set to evaluate membership stability across alternative weighting schemes, and the percentage of the baseline top-20% priority set retained under each alternative weighting was calculated. Robustness was summarized for a moderate abundance-weight range of 0.40-0.60 and a broader plausible range of 0.20-0.80. County-specific membership of the baseline top-20% group across alternative weights was additionally examined.

A separate sensitivity analysis was conducted to evaluate the influence of reported tick-bite exposure data on the vector-level risk scores. For counties with tick-bite incidence, the sensitivity score was calculated as an equal-weight additive combination of tick abundance, pathogen prevalence, and tick-bite incidence:

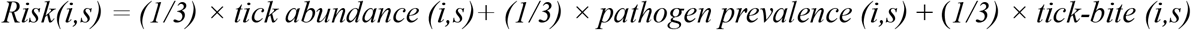

For counties with missing tick-bite incidence, the sensitivity score was calculated using the same equal-weight combination of tick abundance and pathogen prevalence as in the primary analysis. The sensitivity-analysis scores were compared with the primary risk scores using Spearman’s rank correlation to assess the stability of county-level rankings after incorporating tick-bite exposure.

### Validation of risk scores

The epidemiological relevance of the primary risk scores was assessed using negative binomial regression. Negative binomial regression models were fitted using the *glm*.*nb* function in the **MASS** package in R (*R: The R Project for Statistical Computing*, n.d.; Ripley & Venables, 2026) to model county-level cumulative TBD case counts as a function of the corresponding scaled risk score, with log(population) included as an offset to account for county population size:

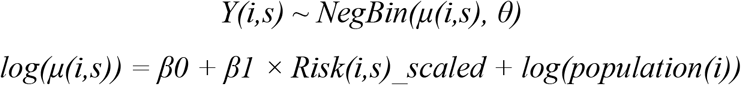

where Y(i,s) is the observed cumulative case count, μ(i,s) is the expected count, and θ is the negative binomial dispersion parameter. Incidence rate ratios (IRR) and their 95% confidence intervals were reported for each species.

### Seasonality

Monthly seasonal patterns were characterized separately for each tick species using three species-specific indicators: human TBD cases, reported tick-bite encounters, and sampled tick abundance from the IDPH. All three variables were summarized by calendar month, and seasonal patterns were visualized using scatter plots with locally estimated scatterplot smoothing (LOESS) curves to depict nonlinear monthly trends across the study period. A composite phenology indicator was then constructed for each species by summing its monthly TBD cases, tick-bite encounters, and sampled tick abundance:

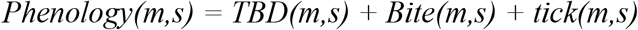

where m denotes calendar month, and s denotes tick species. The resulting species-specific phenology indicators were scaled to represent the relative seasonal activity of each tick species.

To evaluate climatic associations with seasonal activity, statewide daily mean temperature and precipitation data for Illinois were obtained for 2019-2022 (Durre et al., 2022). A 365-day climatology was generated by averaging daily values across the four study years, excluding February 29. Daily climate values were then summarized by calendar month as mean temperature and total precipitation. One-month-lagged climate variables were generated, with December conditions assigned as the preceding-month values for January, to explore whether phenology in a given month is associated with climate in that month or in the preceding month.

Segmented regression models were fitted using the **segmented** package in R (Muggeo, 2003) to identify potential breakpoints in relationships between phenology and concurrent climatic conditions. Models with a single breakpoint were fitted to temperature and precipitation for each tick species.

### Seasonal risk

Species-specific monthly seasonal risk was estimated by combining each county’s vector-level risk score with the corresponding monthly phenology indicator. For county *i*, month m, and species *s*, seasonal risk was calculated as follows:

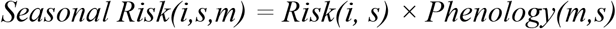

where *Risk(i,s)* represents the county-level vector risk for species *s*, and *Phenology(m,s)* represents the relative seasonal activity of that species during the month m.

To characterize broader spatial patterns, monthly seasonal-risk values were spatially smoothed using a row-standardized, first-order queen-contiguity spatial weights matrix, in which counties sharing a boundary or vertex were considered neighbors (Supplementary materials). Interactive species-specific maps were subsequently incorporated into an R Shiny application deployed on shinyapps.io, allowing users to select individual months and view the county, tick species, the monthly seasonal risk value, and the corresponding risk category.

## Results

### Spatial distribution of risk-score components

County-level data were successfully assembled and linked to the Illinois county boundary file for all 102 counties. Predicted tick abundance and population data were available statewide for all three tick species (Figure 2). Pathogen-testing coverage varied across species and counties, resulting in unstable or missing raw prevalence estimates in counties with limited or no testing (Figure S1). Spatial smoothing generated pathogen-prevalence estimates for all 102 counties (Figure 2). Tick-bite encounter data were available for 37 counties and were therefore used only in sensitivity analyses rather than in the primary vector-level risk score (Figure S2)

**Figure 2.**
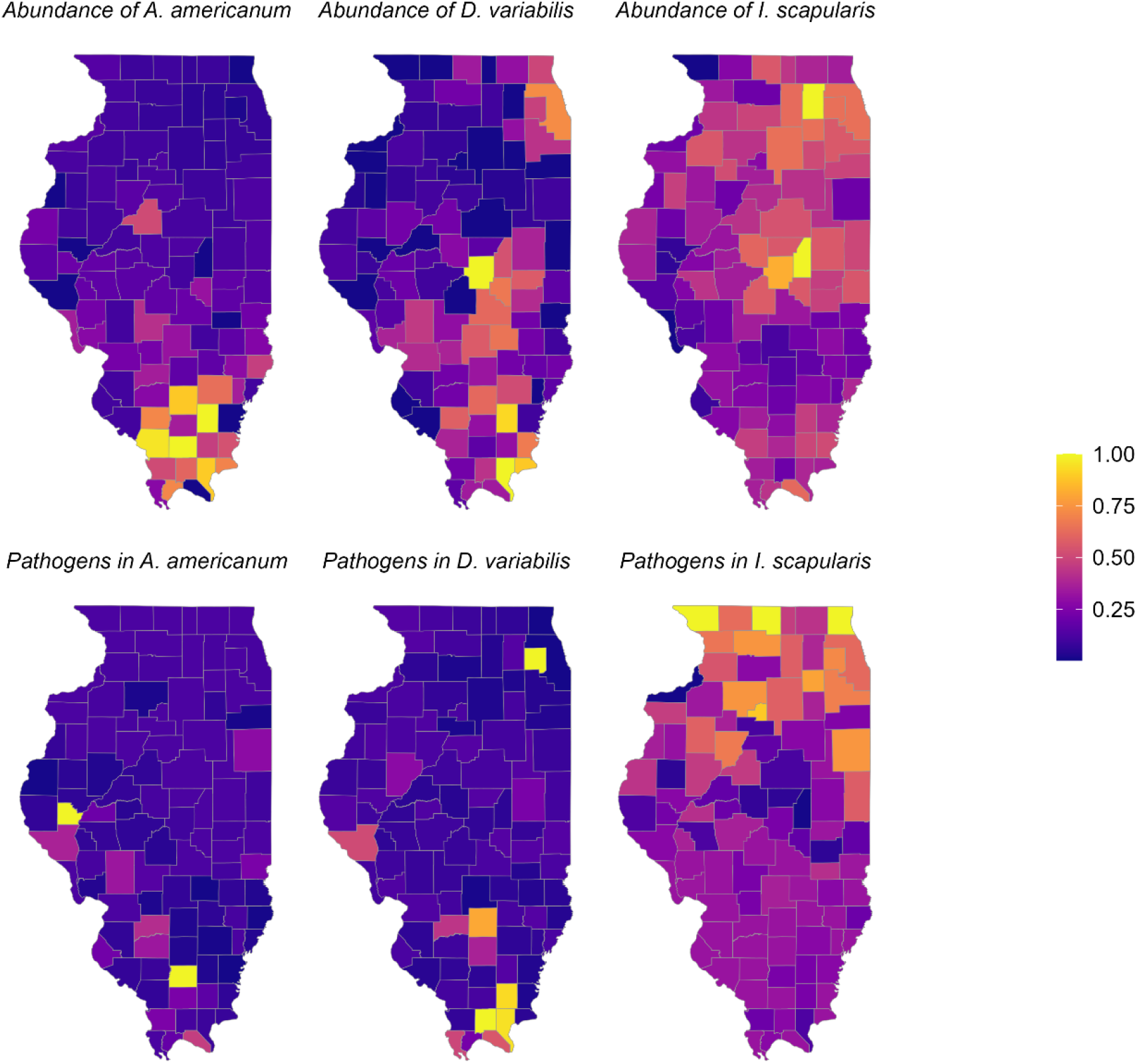
Spatial distribution of scaled predicted tick abundance and scaled smoothed pathogen prevalence for three medically important tick species across Illinois counties. Upper panels show predicted tick abundance and lower panels show smoothed pathogen prevalence, with higher values indicating greater relative abundance or prevalence within each species.

### Risk scores and sensitivity analyses

The primary vector-level risk scores showed substantial geographic variation across Illinois and distinct spatial patterns among the three tick species (Figure 3). The *A. americanum* risk surface was characterized by relatively higher scores in southern Illinois, reflecting the concentration of predicted abundance and elevated pathogen prevalence in this region. Risk for *D. variabilis* was more spatially heterogeneous, with elevated scores distributed across several southern, central, and eastern counties. The *I. scapularis* risk surface showed a broader distribution of moderate-to-high scores, including counties in northern, central, and eastern Illinois. Because the scores were scaled separately for each species, values represent relative county-level risk within a species and should not be used to compare the absolute magnitude of risk among species.

**Figure 3.**
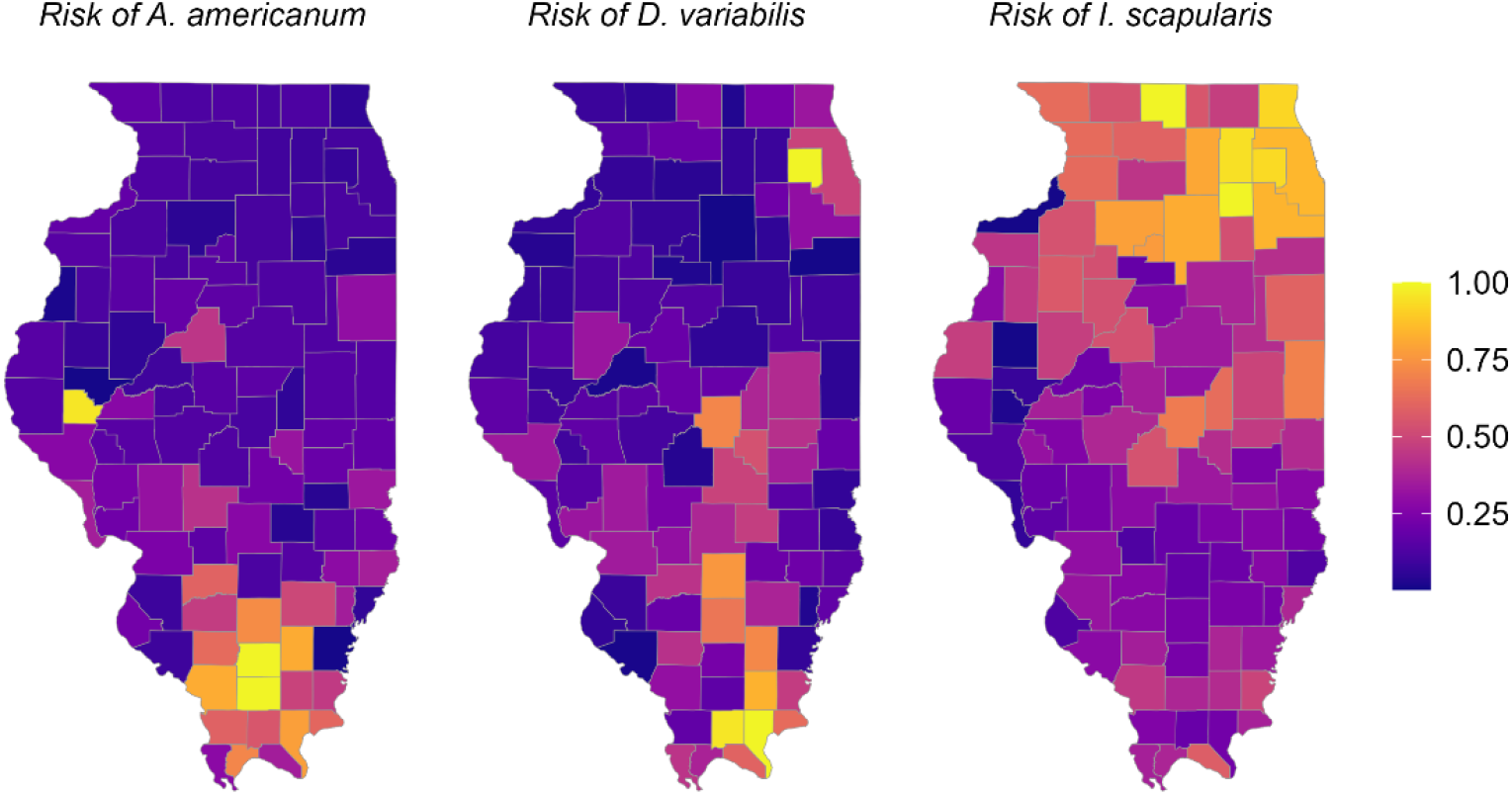
Spatial distribution of species-specific vector-level risk scores of three medically important tick species across Illinois counties.

Deterministic weight sensitivity analysis indicated that county rankings were highly stable around the equal-weight baseline (Figure 4). Across abundance weights of 0.40–0.60, the minimum Spearman correlations with the 50:50 baseline were 0.982 for *A. americanum*, 0.989 for *D. variabilis*, and 0.980 for *I. scapularis*. At least 90.5% of the 21 counties in the baseline top-20% priority set were retained across this range for all three species. Across the broader abundance-weight range of 0.20–0.80, minimum Spearman correlations remained 0.863, 0.892, and 0.836, respectively, while minimum retention of the baseline top-20% priority set was 71.4%, 71.4%, and 66.7%, respectively. Thus, the broad statewide ranking structure was relatively robust to alternative abundance-prevalence weighting, although membership near the high-risk threshold became more sensitive as weights moved farther from the equal-weight formulation. County-specific stability of the baseline high-risk group across the weight range is shown in the supplementary files (Figure S3).

**Figure 4.**
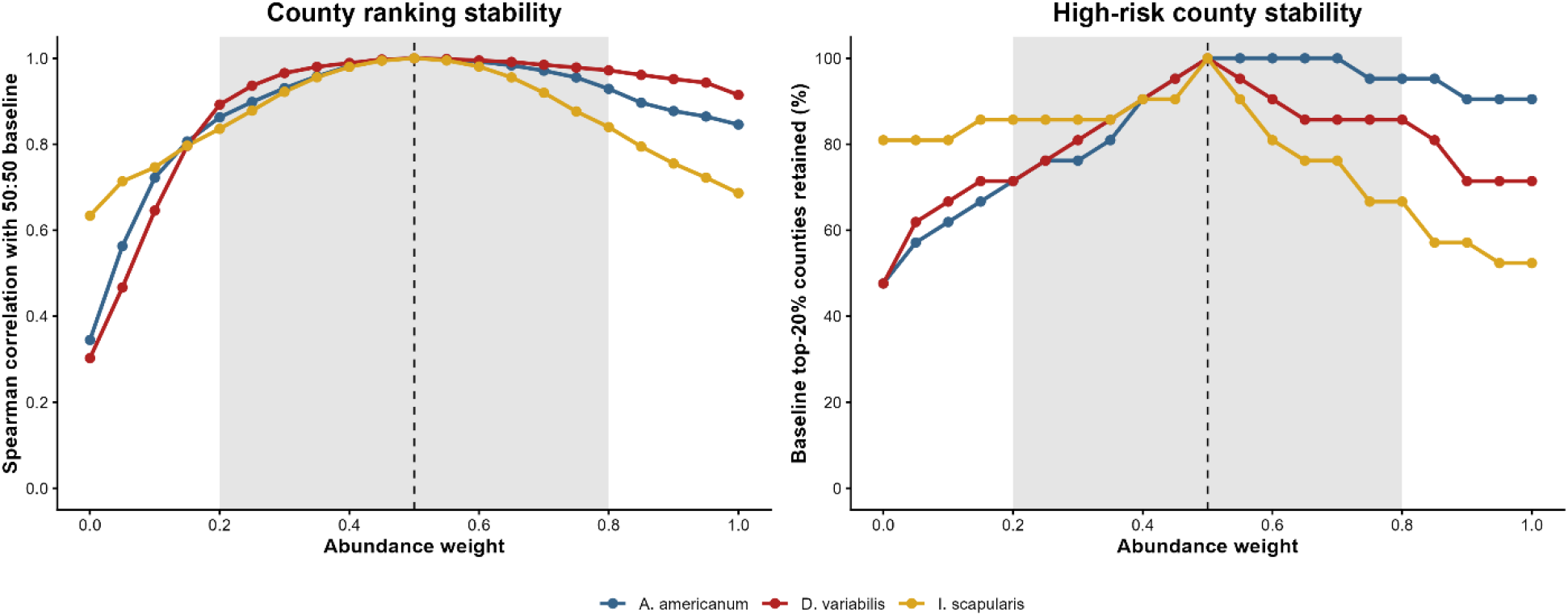
Deterministic sensitivity of county-level risk rankings to alternative abundance-prevalence weights. Left: Spearman correlation between alternative-weight rankings and the primary 50:50 formulation. Right: percentage of the baseline top-20% priority set retained across alternative weights. The shaded region indicates abundance weights of 0.20–0.80, and the dashed line marks the 0.50 baseline.

In the separate exposure sensitivity analysis, including reported tick-bite incidence, produced risk-score rankings that were strongly correlated with those from the primary analysis. Spearman’s rank correlations between the primary and sensitivity scores were 0.952 for *A. americanum*, 0.961 for *D. variabilis*, and 0.882 for *I. scapularis*; all correlations were statistically significant. These findings indicated that county rankings were generally robust to the inclusion of reported tick-bite exposure, although the comparatively lower correlation for *I. scapularis* suggested greater changes in its county-level rankings (Figure 5).

**Figure 5.**
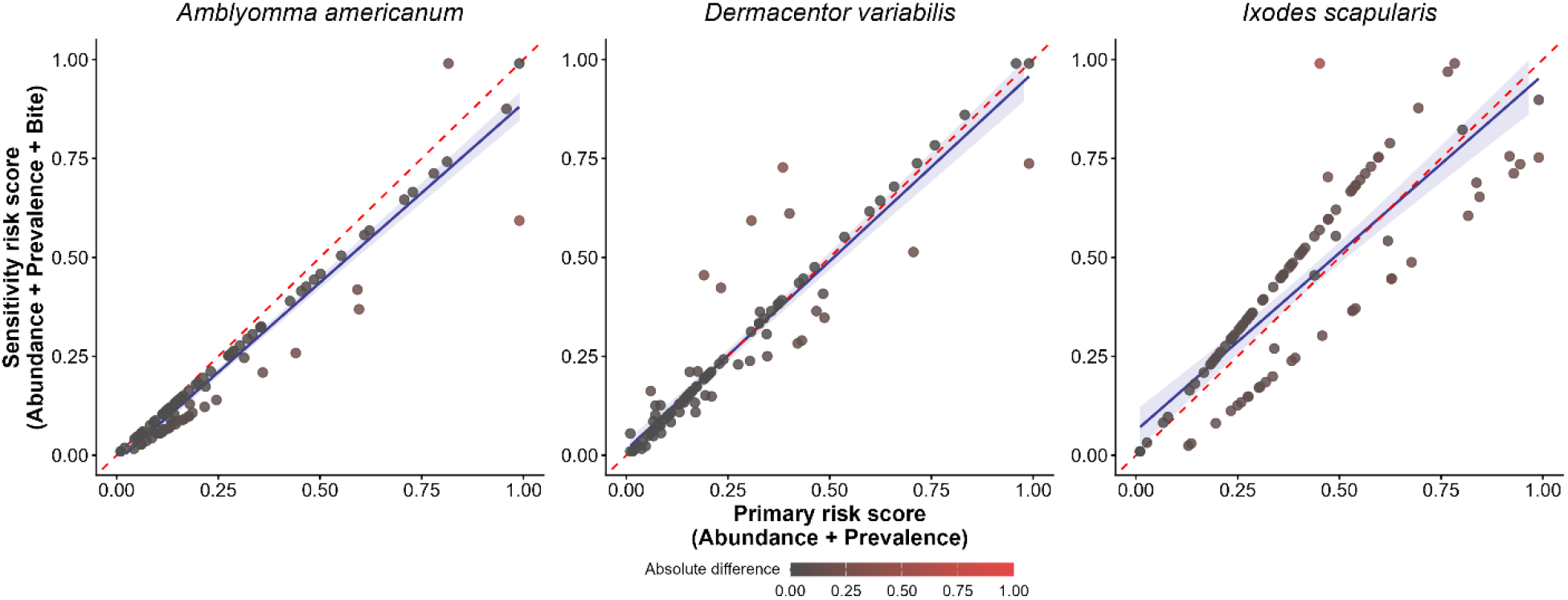
Agreement between primary risk scores and tick-bite-informed sensitivity scores across Illinois counties. Scatter plots compare the primary risk score, based on predicted tick abundance and pathogen prevalence, with the sensitivity score, which incorporates reported tick-bite incidence where available. The red dashed line represents perfect agreement between the two scores, while the blue solid line represents the fitted linear relationship and the shaded band indicates its 95% confidence interval. Point color represents the absolute difference between the primary and sensitivity scores.

### Validation of risk score

Negative binomial regression indicated that higher vector-level risk scores were associated with greater county-level TBD case counts for all three species after accounting for county population (Figure S4). Effect estimates were expressed as incidence rate ratios for a 0.2-unit increase in the scaled risk score, calculated as exp(0.2β_1_), with 95% confidence intervals transformed on the same scale. For each 0.2-unit increase in the scaled risk score, the expected case rate increased by 2.76-fold for *A. americanum* (IRR = 2.76, 95% CI: 2.13-3.67; P <0.001), 2.44-fold for *D. variabilis* (IRR = 2.44, 95% CI: 1.65-3.85; P <0.001), and 1.47-fold for *I. scapularis* (IRR = 1.47, 95% CI: 1.20-1.82; P <0.001) (Figure 6).

**Figure 6.**
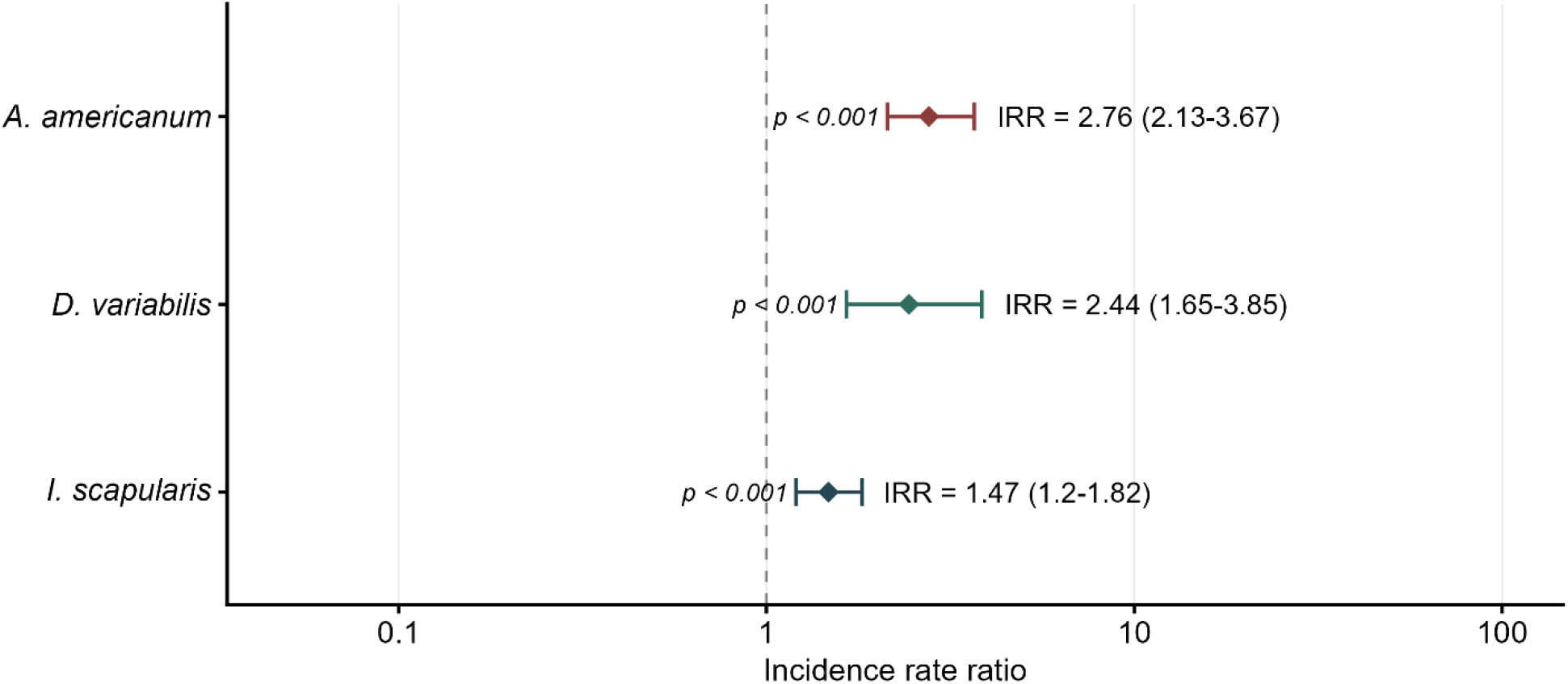
Association between species-specific vector-level risk scores and human tick-borne disease rates across Illinois counties. Diamonds represent incidence rate ratios (IRRs) for each 0.2-unit increase in the scaled vector-level risk score from species-specific negative binomial regression models with county population included as an offset. Horizontal lines indicate 95% confidence intervals; IRRs above 1 indicate increasing disease rates with increasing risk scores, values below 1 indicate decreasing rates, and the dashed line at 1 indicates no association.

### Seasonal patterns and climatic associations

A 365-day climatology was generated by averaging daily values across the four study years (Figure S5). The composite phenology indicators showed clear seasonal variation for all three tick species (Figure 7). Seasonal activity was low in winter, increased in spring, reached its highest levels in late spring and summer, and then declined toward the end of the year. *A. americanum* and *D. variabilis* exhibited pronounced warm-season patterns, whereas *I. scapularis* showed a broader seasonal profile extending across a greater portion of the year.

**Figure 7.**
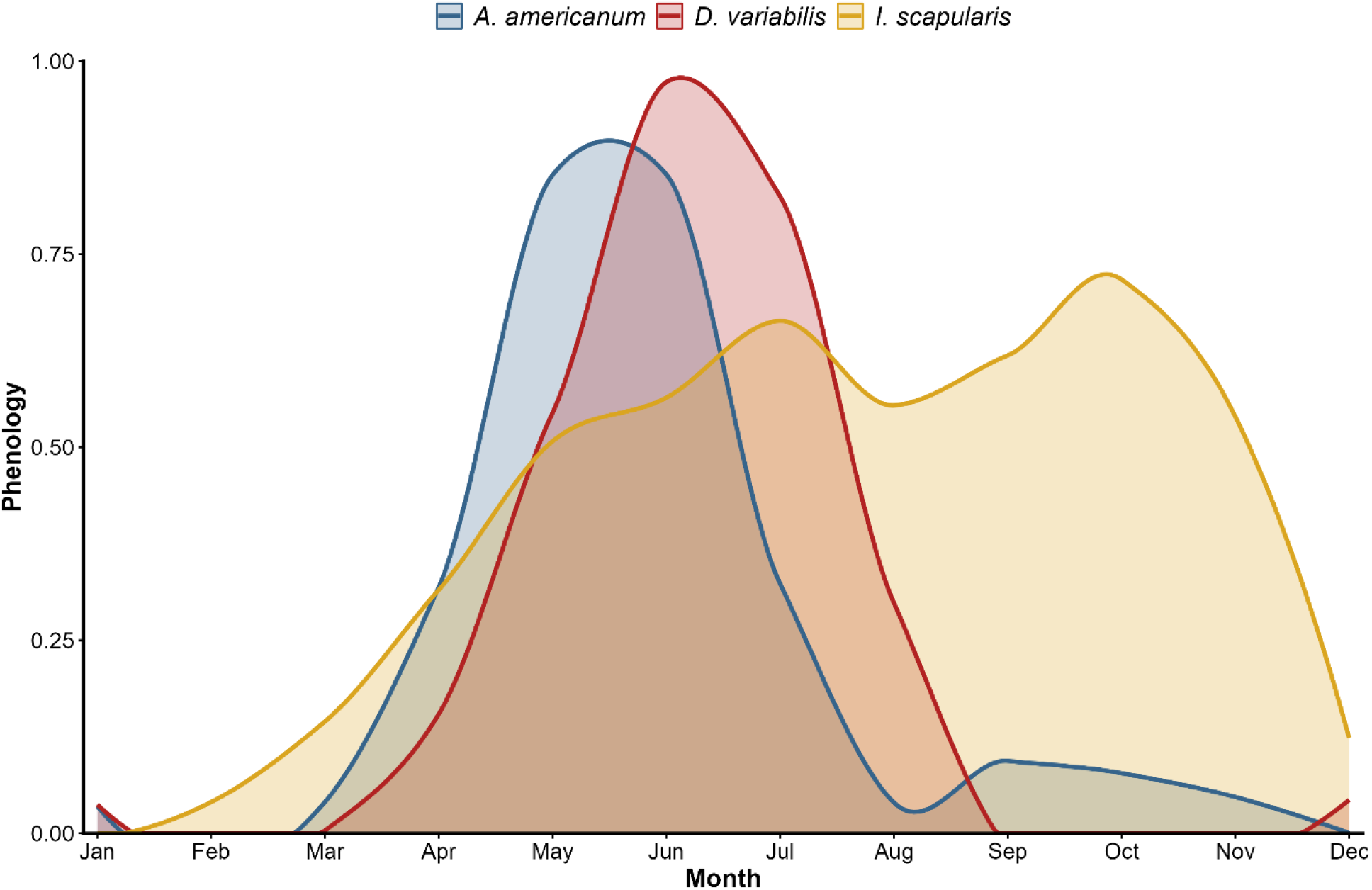
Species-specific monthly phenology patterns for three medically important tick species in Illinois. Phenology indicators were constructed by combining monthly human tick-borne disease cases, reported tick-bite encounters, and sampled tick abundance for medically important tick species.

The mean monthly temperature was positively correlated with phenology for all three species. Temperature was positively associated with phenology for *I. scapularis* (*ρ*= 0.748, *p*= 0.007) and *D. variabilis* (*ρ*= 0.587, *p*= 0.049), whereas the association for *A. americanum* did not reach statistical significance (*ρ*= 0.573, *p*= 0.055). While precipitation was positively associated with phenology for all three species and showed the strongest association for *D*.

*variabilis* (*ρ* = 0.839, *p* = 0.001), followed by *A. americanum* (*ρ* = 0.720, *p* = 0.011) and *I. scapularis* (*ρ*= 0.699, p= 0.015).

Segmented regression identified changes in the relationships between phenology and climatic conditions (Figure 8). Estimated temperature breakpoints occurred at approximately 22°C for *A. americanum*, 12.8°C for *D. variabilis*, and 12.7°C for *I. scapularis*. Phenology generally increased with temperature below these thresholds, although the response differed among species after the breakpoint. For *D. variabilis*, phenology increased more rapidly above the breakpoint, whereas *I. scapularis* showed a slight decline in the slope beyond approximately 13°C. In contrast, *A. americanum* exhibited a reduced response at the highest temperatures. Estimated precipitation breakpoints occurred at approximately 93.6 mm for *A. americanum*, 79.9 mm for *D. variabilis*, and 54.4 mm for *I. scapularis*. Phenology generally increased with precipitation, although the rate of increase differed before and after the estimated thresholds. *A. americanum* and *D. variabilis* exhibited steeper positive relationships beyond their respective breakpoints, whereas *I. scapularis* showed a more gradual increase across the observed precipitation range. Because these segmented models were based on only 12 monthly observations, the breakpoint estimates should be interpreted cautiously.

**Figure 8.**
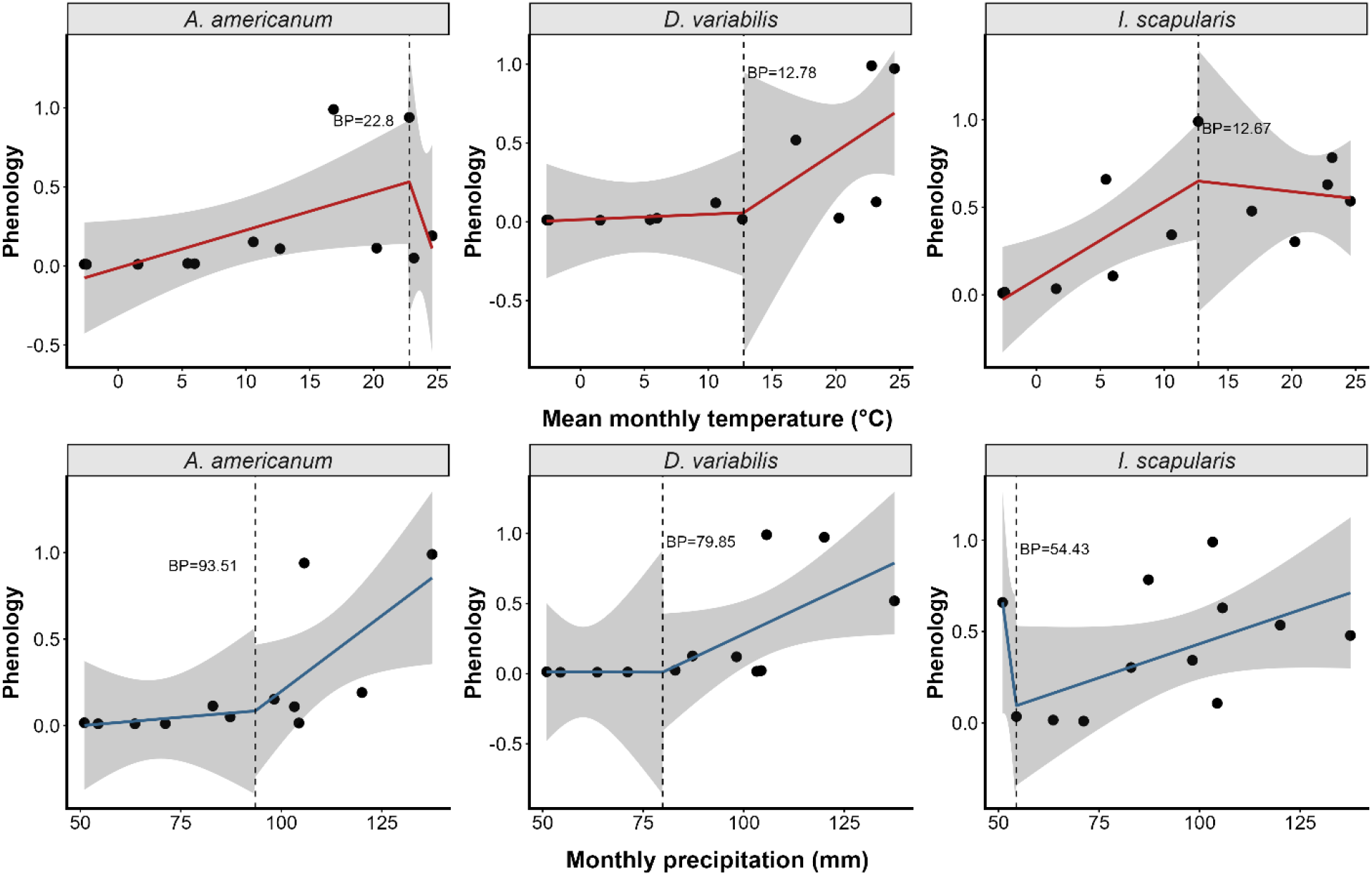
Segmented relationships between species-specific phenology and climatic conditions in Illinois. Panels show relationships between scaled monthly phenology and mean monthly temperature or total monthly precipitation for three medically important tick species. Points represent monthly observations, solid lines represent segmented regression predictions, shaded bands indicate 95% confidence intervals, and dashed vertical lines indicate estimated breakpoints (BP).

### Seasonal risk

Monthly seasonal-risk scores were generated for every county and tick species by multiplying the county’s scaled vector-level risk score by the corresponding species-specific scaled phenology value for each calendar month. The resulting scores showed distinct species-specific seasonal patterns across Illinois (Figure 9). Seasonal risk for *A. americanum* increased during spring, was highest during late spring and early summer, and declined substantially by late summer. *Dermacentor variabilis* showed its highest seasonal risk during early and mid-summer. In contrast, *I. scapularis* exhibited a broader seasonal-risk period extending from spring through autumn, with relatively elevated risk persisting later in the year. The county-by-month calendar heat maps demonstrated substantial geographic and temporal heterogeneity in seasonal risk (Figure 9). Counties were arranged by southern, central, and northern Illinois regions, allowing regional patterns to be compared across the 12 calendar months.

**Figure 9.**
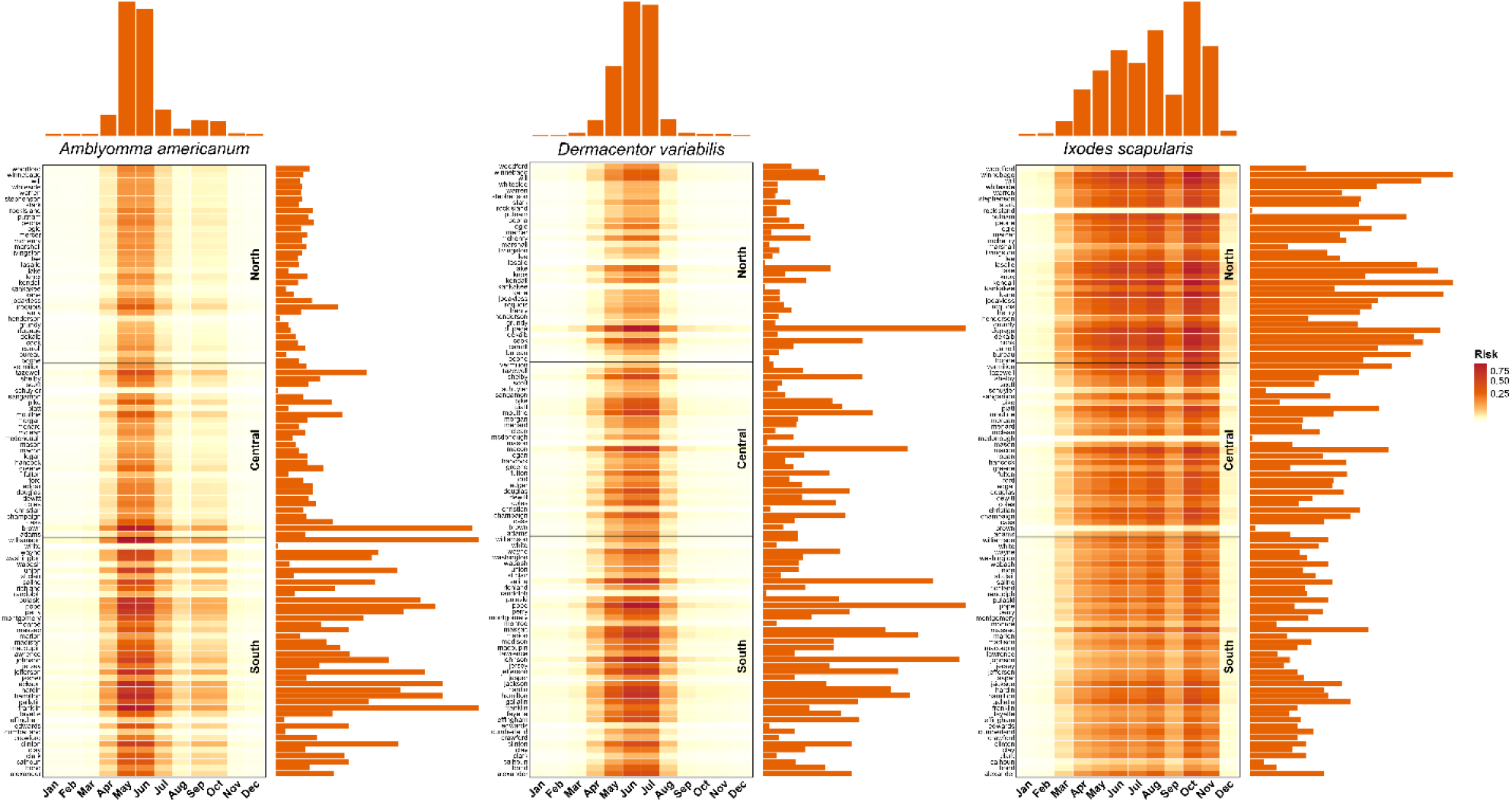
County-level monthly seasonal-risk patterns for three medically important tick species across Illinois. Each row represents an Illinois county, and each column represents a calendar month. Counties are grouped into southern, central, and northern Illinois regions, with horizontal lines separating regions. Cell color indicates relative seasonal risk, while the top and right marginal bars summarize mean risk across months and counties, respectively.

To reduce abrupt differences between adjacent counties and emphasize broader spatial patterns, the monthly seasonal-risk surfaces were spatially smoothed separately for each species and month. Smoothed scores were then assigned to five categories: very low, low, medium, high, and very high. The spatially smoothed monthly risk surfaces were incorporated into an interactive R Shiny application (https://abrarhussain.shinyapps.io/interactive_maps). The application allowed users to select a tick species and a calendar month, then view the corresponding county-level seasonal risk distribution across Illinois. Selecting a county displayed its name, species, smoothed seasonal-risk value, and assigned risk category. This interactive format provided a month-specific visualization of changes in the geographic distribution and intensity of seasonal risk throughout the year.

## Discussion

This study developed a species-specific framework for characterizing tick-borne disease risk across Illinois by integrating predicted tick abundance and pathogen prevalence, evaluating human exposure and disease burden, and incorporating seasonal activity. The central value of the framework is that it treats these components as related but distinct stages in the pathway from vector presence to human disease. Tick abundance reflects the opportunity for encounter; pathogen prevalence reflects the probability that encountered ticks are infected; human exposure influences whether ecological hazard translates into contact; and seasonal activity determines when these processes are most likely to overlap. This integrated perspective is consistent with earlier acarological risk approaches, which demonstrated that combining vector abundance with infection prevalence is more informative than considering either component alone (Diuk-Wasser et al., 2012; Mather et al., 1996; Pepin et al., 2012).

The geographic structure of the resulting risk surfaces was also ecologically plausible. Higher *A. americanum* risk in southern Illinois is consistent with previous surveillance showing stronger abundance and clustering of this species in the southern part of the state (Hussain et al., 2025), *D. variabilis* showed a broader and more heterogeneous distribution, while *I. scapularis* showed substantial risk across northern and central Illinois. Similar spatial differentiation among these species has been reported elsewhere in the Midwest, where climate, forest structure, host availability, and proximity to established populations shape tick occurrence and expansion (Gardner et al., 2020; Johnson et al., 2018). These patterns are important because they reinforce that statewide risk is not spatially uniform and that different vector species may require different geographic priorities for surveillance and prevention.

The present framework follows the same conceptual logic as classical measures, such as the density of infected nymphs, which combine tick abundance and pathogen infection to represent the entomological hazard. Mather et al. showed that an entomological index based on the prevalence of infected *I. scapularis* was associated with the risk of human Lyme disease (Mather et al., 1996). Diuk Wasser et al. later used standardized field sampling to map the distribution of infected host-seeking nymphs across the eastern United States (Diuk-Wasser et al., 2012), and Pepin et al. reported positive associations between infected nymph density and county-level Lyme disease incidence (Pepin et al., 2012). The current approach differs in an important way. Rather than estimating the absolute density of infected ticks, abundance and pathogen prevalence were scaled separately and combined using equal weights. The resulting score should therefore be interpreted as a relative prioritization index rather than a direct probability of infection. The overall structure is also conceptually consistent with CDC surveillance priorities that integrate information on vector distribution and abundance, pathogen prevalence, and seasonal activity (CDC, 2019, 2020). This is especially relevant because the framework was designed for three tick species with different ecologies and associated disease groups. Equal scaling provides a common structure across species, but the scores are best interpreted within species rather than as absolute comparisons among species.

The deterministic sensitivity analysis further showed that the broad county-priority structure was not strongly dependent on the exact choice of 50:50 weighting. Rankings remained highly concordant under moderate changes in abundance and prevalence weights, and substantial agreement persisted across the broader 0.20-0.80 range of abundance weights. However, declining retention of the baseline high-risk group toward the boundaries of this range indicates that counties near the prioritization threshold are more sensitive to the weighting assumption than the overall statewide ranking.

Integrated risk mapping studies have also shown that ecological hazard does not always align perfectly with human disease because exposure depends on human behavior and contact with suitable habitat. Bouchard et al. demonstrated that ecological and social-behavioral information can identify distinct geographic priorities for Lyme disease prevention (Bouchard et al., 2018). Similarly, Eisen and Eisen emphasized that tick-based measures may correlate well with disease at broader spatial scales while still failing to capture all variation in human exposure (L. Eisen & Eisen, 2016). The separate tick-bite sensitivity analysis supports that interpretation, as the inclusion of reported tick-bite incidence altered some county rankings while leaving the overall spatial structure largely intact.

The positive association between vector-level risk and human tick-borne disease case rates supports the score’s epidemiological relevance. This is consistent with previous studies showing that greater acarological hazard is associated with higher human disease incidence (Diuk-Wasser et al., 2012; Mather et al., 1996; Pepin et al., 2012). By modeling reported case counts while accounting for county population, the negative binomial models preserved variation in disease burden and allowed evaluation of whether increasing vector-level risk corresponded to increasing human disease rates. The strength of this association differed among the three vector-associated disease groups, indicating that ecological hazard may translate into reported human disease differently among tick species and their associated pathogens. Such differences may reflect variation in human exposure, healthcare-seeking behavior, diagnostic practices, reporting completeness, and the fact that county of residence may not always correspond to the actual location of infection. Thus, although vector-level risk was positively associated with human disease burden, ecological hazard represents only one component of observed disease occurrence, with behavioral, healthcare, and surveillance processes also influencing the number of reported cases.

The tick-bite sensitivity analysis showed that adding reported tick-bite incidence generally preserved county rankings, suggesting that the abundance and pathogen components captured much of the large-scale geographic pattern of risk. However, the somewhat lower agreement for *I. scapularis* suggests that ecological hazard and reported exposure may diverge in some areas. This distinction is supported by citizen science studies showing that human-submitted tick records can provide information that differs from conventional environmental surveillance. Nieto et al. demonstrated that voluntary tick submissions captured broad patterns of human contact with multiple vector species and pathogens across the United States (Nieto et al., 2018). Exposure data, therefore, add an important behavioral dimension to ecological risk, but they can also be affected by uneven participation and reporting intensity. Given the limited geographic coverage of reported tick-bite encounters in the current study, using them as a sensitivity component rather than as a core part of the primary score was a reasonable strategy.

The phenology results reinforce the importance of treating tick-borne disease risk as dynamic rather than static. *A. americanum* and *D. variabilis* showed concentrated warm-season activity, whereas *I. scapularis* displayed a broader seasonal profile. These patterns are consistent with field observations from the Midwest. Studies in Illinois and neighboring states have documented strong spring and summer activity for *A. americanum* and *D. variabilis*, while *I. scapularis* commonly shows activity extending into cooler periods, particularly among adults (Burtis et al., 2023; Gilliam et al., 2018; Hamer et al., 2012). The broader seasonal pattern of *I. scapularis* is also consistent with its life history. Early stages are generally most active during warmer months, while adults may show spring and fall peaks (Burtis et al., 2023; Hamer et al., 2012). A composite phenology indicator integrating sampled ticks, bite encounters, and human disease would therefore be expected to show a wider seasonal window than a measure based on a single life stage.

Temperature and precipitation associations further support the biological basis of these patterns. Temperature influences development and host-seeking behavior, while moisture availability affects survival and desiccation risk. Climate-based studies have shown that both temperature and precipitation help determine the distribution and activity of *I. scapularis* (Brownstein et al., 2003). Similar relationships have been reported for *A. americanum*, where seasonal activity responds to weather conditions and habitat characteristics (Gilliam et al., 2018). The positive associations with preceding month precipitation in the present analysis are also biologically plausible because moisture conditions can influence survival and activity over periods longer than a single sampling day. The segmented relationships suggest that climatic effects may not be linear across the observed range. Increasing temperature may enhance activity under cool conditions but provide diminishing benefits at higher temperatures, particularly if desiccation risk increases. Likewise, additional precipitation may be most influential when moisture is limited. Comparable geographic variation in phenology has been reported for *I. scapularis* across the United States (Ogden et al., 2018). Because the present breakpoint estimates were based on only 12 average monthly observations of the study period, they should be interpreted as exploratory rather than as fixed physiological thresholds.

Combining annual county-level ecological risk with species-specific phenology provides a more actionable interpretation of when elevated risk is likely to occur. Static annual maps identify geographic priorities, but they do not indicate when those priorities become most relevant. The seasonal risk framework adds this temporal dimension by showing that the timing and duration of elevated risk differ among species. This has practical implications for surveillance and communication. Prevention messaging for *A. americanum* and *D. variabilis* may be most relevant during concentrated warm-season periods, whereas *I. scapularis* may require a broader communication window that extends into cooler months. Similar conclusions have emerged from mechanistic models showing that temperature-driven changes in tick activity can shift the timing and duration of Lyme disease risk (Li et al., 2016).

The interactive R Shiny application extends the analytical framework by allowing users to examine month-specific county-level risk rather than relying on a single annual map. This may improve the usefulness of the results for local health departments, clinicians, researchers, and the public by linking spatial prioritization with seasonal timing.

Several limitations should be considered. The underlying datasets differed in surveillance intensity and completeness. Pathogen testing varied across counties and species, tick-bite reports were geographically incomplete, and human disease surveillance was influenced by diagnostic and reporting practices. Spatial smoothing reduces instability in sparse prevalence estimates but may also attenuate true local extremes. County-level analysis can also obscure finer-scale variation in tick and pathogen occurrence, host communities, human behavior, and local landscape characteristics such as land cover, vegetation structure, habitat patch size, and fragmentation. Finer-scale surveillance and environmental data could therefore improve identification of localized areas of elevated risk within counties. The equal weighting strategy was selected for transparency and comparability and should not be interpreted as evidence that abundance and pathogen prevalence contribute equally to transmission in biological terms. Although the deterministic sensitivity analysis showed that broad county rankings were robust across a range of alternative weights, the classification of some counties near the high-risk threshold remained sensitive to the choice of weights. The seasonal framework could also be improved by estimating regional or county-specific phenology as longer series become available.

## Conclusion

Overall, this study supports a view of tick-borne disease risk as the interaction of ecological hazard, pathogen presence, human exposure, disease occurrence, and seasonality. The framework builds on established acarological risk concepts while extending them to three medically important tick species within a common statewide system. Its main strength is not the production of a single definitive probability of infection, but the identification of relative geographic and seasonal priorities that can support surveillance, prevention, and future data collection in Illinois.

## Acknowledgments

We sincerely thank the researchers and surveillance personnel who contributed to documenting tick occurrence records in Illinois. We also gratefully acknowledge the Illinois Department of Public Health for providing the data used in this study.

## CRediT authorship contribution statement

**Abrar Hussain:** Conceptualization, Methodology, Data Curation, Visualization, Formal analysis, Software, Writing-original draft, Writing-review & editing; **Suman Bhowmick**: Formal analysis, Writing-review & editing; **Nohra Mateus-Pinilla**: Writing-review & editing; **Rebecca L. Smith**: Conceptualization, Writing-review & editing, Supervision.

## Data Availability Statement

Data and analysis code used in this study are publicly available at: https://github.com/abrarhussain95/IL_TBD_Risk

