## Supplementary file for "Mapping county-level tick-borne disease risk in Illinois by integrating vector abundance, pathogen prevalence, and phenology"

***Supporting Information***

Supplementary File S1. Supplementary methods and figures. This file provides additional methodological details on pathogen prevalence smoothing, scaling of risk-score components, and spatial smoothing of monthly seasonal risk. Five supplementary figures present unsmoothed pathogen prevalence (Figure S1), reported tick-bite counts (Figure S2), stability of the top-20% county priority set across alternative weighting schemes (Figure S3), reported human tick-borne disease case counts (Figure S4), and statewide daily temperature and precipitation climatology (Figure S5).

### Materials and Methods

***Pathogen prevalence smoothing***

For each tick species, county-level pathogen prevalence was locally smoothed using first-order queen-contiguous neighboring counties. For counties with observed prevalence, the county estimate was combined with the mean prevalence of neighboring counties, with greater weight given to the observed county estimate when more ticks had been tested. For counties without an observed prevalence estimate, the mean prevalence of neighboring counties with available data was used. This procedure was performed separately for each tick species to preserve local spatial variation in pathogen prevalence.

***Scaling***

To place variables measured on different scales onto a comparable range, predicted tick abundance, adjusted pathogen prevalence, and tick-bite incidence were scaled separately for *A. americanum*, *D. variabilis*, and *I. scapularis*. For each variable, values below the 1st percentile were replaced by the 1st-percentile value, and values above the 99th percentile were replaced by the 99th-percentile value to limit the influence of extreme observations. The resulting values were then minmax transformed and shifted to a range of 0.01–0.99. Tick-bite incidence was scaled with zero values included in the transformation.

***Seasonal risk***

Spatial smoothing was applied separately to each monthly species-specific seasonal-risk surface using a first-order queen-contiguity spatial weights matrix. Counties sharing either a common boundary or vertex were considered neighbors. The weights were row-standardized so that the weights assigned to all neighboring counties of a given county summed to 1. For each county, species, and month, the smoothed seasonal-risk value was calculated as the weighted mean of the corresponding seasonal-risk values in neighboring counties; the focal county's own value was not included. This procedure was repeated independently for all 12 months and all three tick species, while the original unsmoothed values were retained separately. Smoothed values were classified using fixed thresholds as very low (0–0.01), low (>0.01–0.10), medium (>0.10–0.30), high (>0.30–0.60), and very high (>0.60–1.00), and these categories were used in the interactive monthly risk maps.

### Results

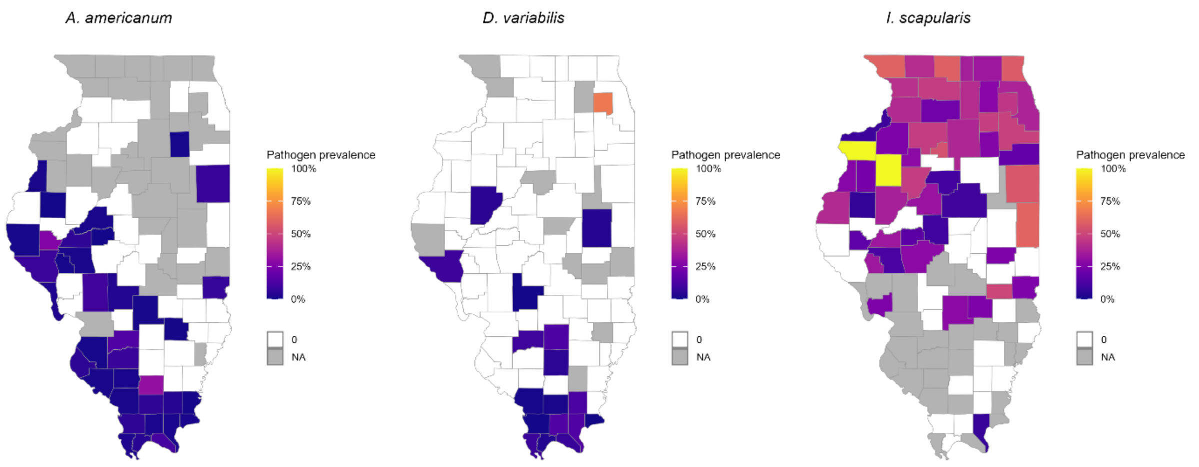


**Figure S1.** Spatial distribution of unsmoothed and unscaled tick-borne pathogen prevalence across Illinois counties. Panels show the observed proportion of tested samples from three medically important tick species in Illinois that are positive for at least one associated pathogen. Prevalence was calculated as the number of pathogen-positive ticks divided by the total number of ticks tested within each county.

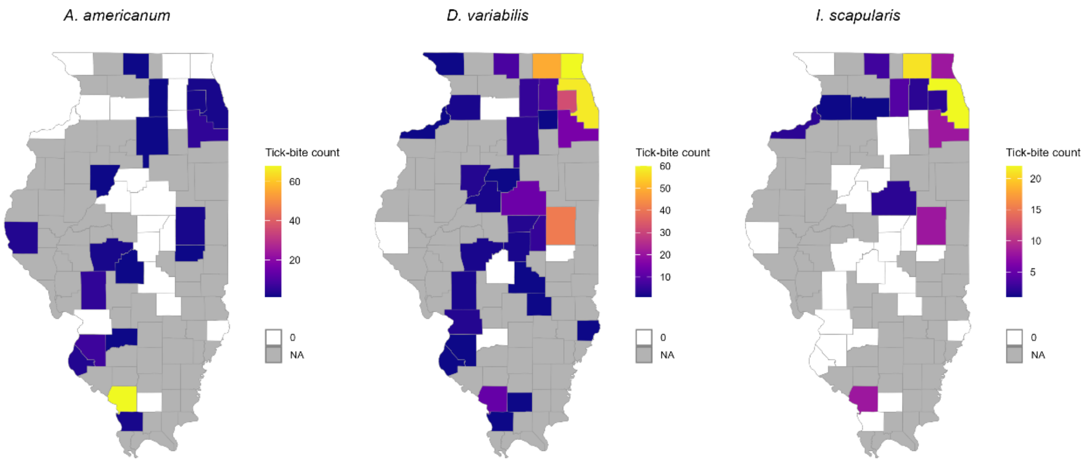


**Figure S2.** Spatial distribution of reported tick-bite counts across Illinois counties. Panels show cumulative reported bite encounters involving three medically important tick species in Illinois.

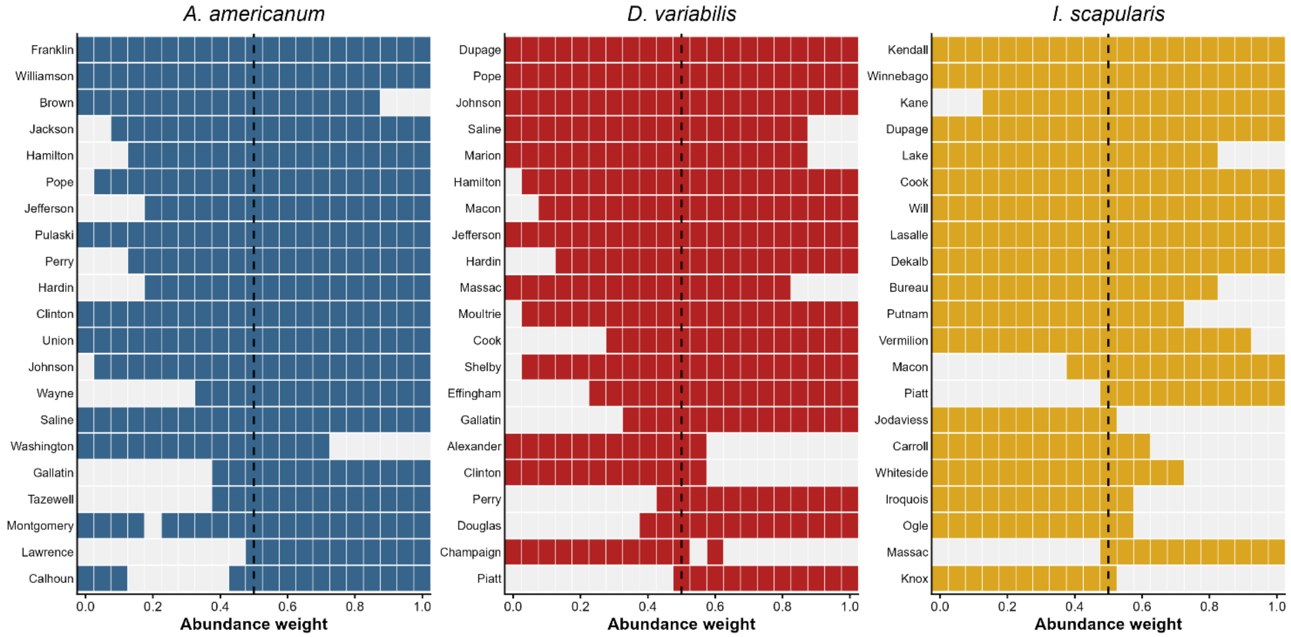


**Figure S3.** Stability of the baseline top-20% county priority set across alternative abundance-prevalence weights. Rows represent the 21 counties in the top 20% under the primary 50:50 formulation. Colored cells indicate that a county remained within the species-specific top 20%, while gray cells indicate movement outside the top 20%. The dashed line marks the primary 0.50 abundance weight.

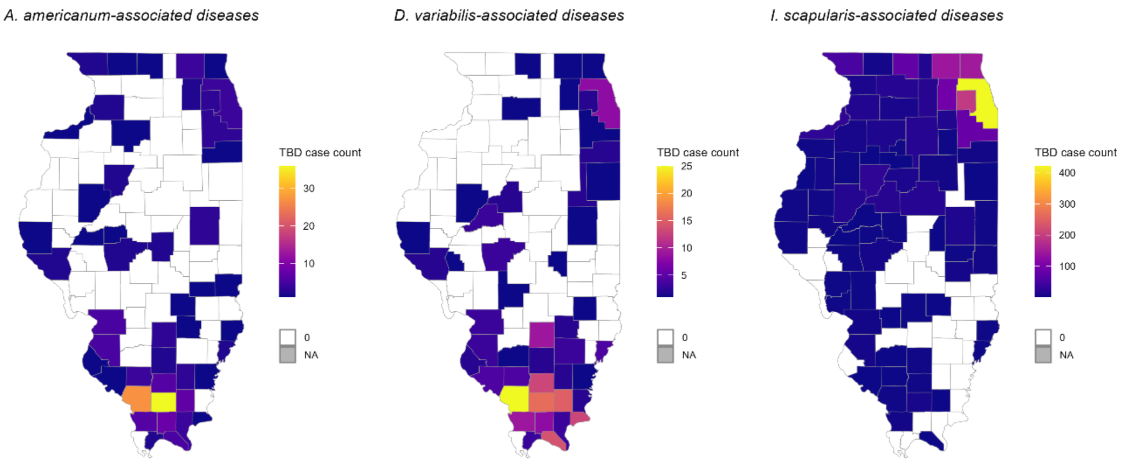


**Figure S4.** Spatial distribution of reported human tick-borne disease case counts across Illinois counties.

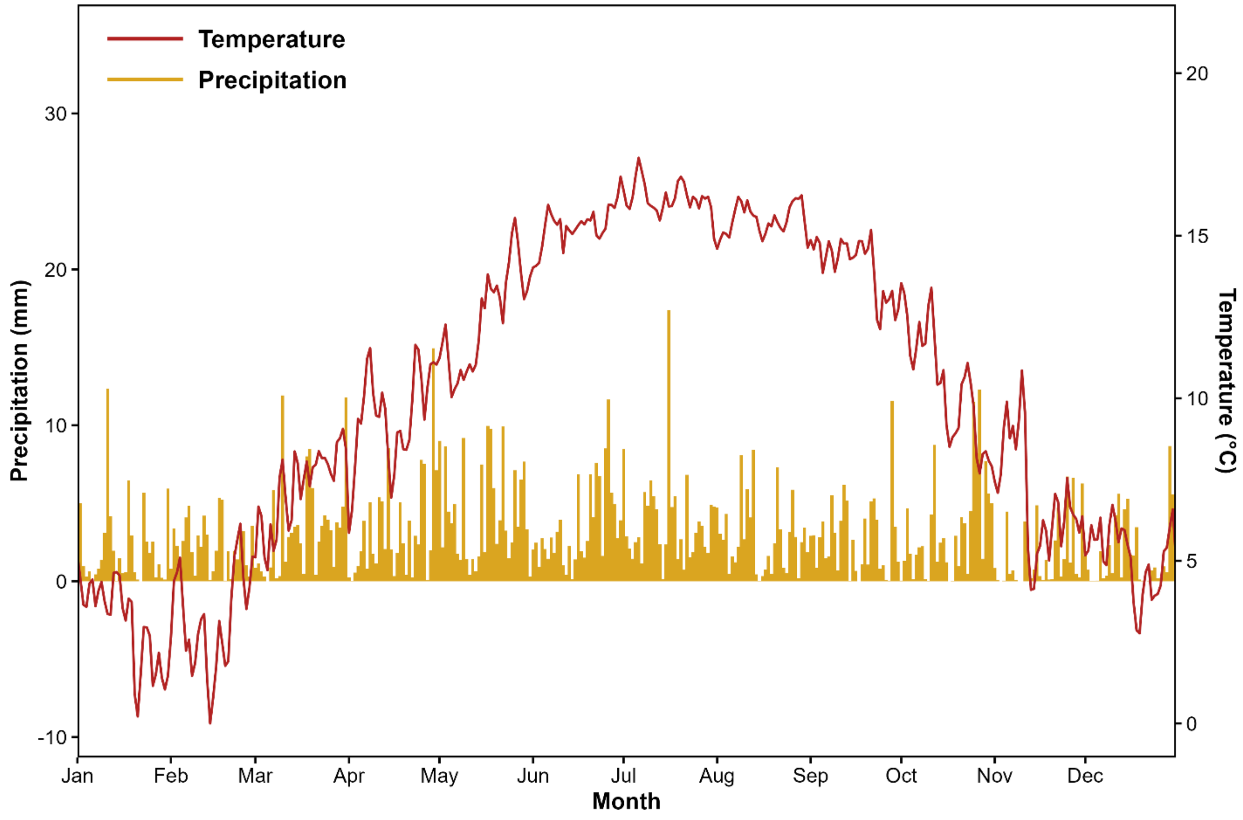

**Figure S5.** Statewide daily climatology of temperature and precipitation in Illinois, 2019–2022. Daily values represent averages across the four study years for each calendar day. The red line shows mean daily temperature (°C; right y-axis), and the gold bars show mean daily precipitation (mm; left y-axis).
